# Genetic testing for *SCA27B* in Korean Multiple System Atrophy

**DOI:** 10.1101/2024.10.21.24315855

**Authors:** Joshua Laß, Michele Berselli, Doug Rioux, Susen Schaake, Jordan Follett, Jonathan E. Bravo, Alexander D. Veit, William Ronchetti, Sarah B. Reiff, Matthew J. Huentelman, Dana Vuzman, Pamela Bower, Peter J. Park, Vikram Khurana, Joanne Trinh, Beomseok Jeon, Han-Joon Kim, Matthew J. Farrer

**Affiliations:** Institute of Neurogenetics, University of Lübeck, Germany; Department of Biomedical Informatics, Harvard Medical School, Boston, MA 02115, USA; Department of Neurology, College of Medicine, University of Florida, Gainesville, FL, USA; Neurogenomics Division, Translational Genomics Research Institute, Phoenix, Arizona, USA; Division of Genetics, Department of Medicine, Brigham and Women’s Hospital, Harvard Medical School, Boston, MA 02115, USA; Mission MSA (formerly MSA Coalition), 1660 International Drive, Suite 600 McLean, VA 22102, USA; Division of Movement Disorders, Department of Neurology, Brigham and Women’s Hospital, Harvard Medical School, Boston, MA 02115, USA; Department of Neurology, Seoul National University Hospital, Seoul National University College of Medicine, Seoul, Korea

## Abstract

*FGF14* (GAA)_n_ repeat expansions are a common cause of idiopathic late-onset ataxia (SCA27B). The cerebellar form of multiple system atrophy (MSA) has comparable clinical features, albeit faster progression. Hence, we performed an analysis of *FGF14* genomic variability in a South Korean cohort of 199 patients with ‘probable’ MSA, compared with 1,048 ethnically-matched controls. All whole genome sequences (WGS) are depicted on a computational genome analysis platform, CGAP, to enable storage, visualization and analysis for partners of the International MSA Coalition. The size of the *FGF14* (GAA)_n_ repeat was also assessed by genomic PCR, and by interrogating WGS data using Expansion Hunter (EH) with an extensive catalogue of potential repeats. However, MSA samples were not significantly different to matched Korean controls, and only three MSA patients showed possible abnormal *FGF14* (GAA)_n_ expansions >300bp. Nevertheless, as PCR and EH findings were often discordant, a subset of samples with expansions was validated by long-read sequencing. Some intermediate expansions (>150 bp) were found in 6.9% (27/392) of controls compared to 13.4% (46/344) in MSA, though overall our results suggest *FGF14* (GAA)_n_ repeat expansions do not influence susceptibility to MSA in Korean patients and highlight challenges inherent in this genetic testing.

## INTRODUCTION

Recent reports suggest *FGF14* (GAA)_n_ expansions (SCA27B) are a common cause of familial and idiopathic late-onset ataxia (ILOCA); (GAA)_n_>335 is disease-causing and fully penetrant, while (GAA)_n_>250 is likely pathogenic with reduced penetrance.^1,2^ ILOCA is a catch-all term that comprises patients who present with ataxia, typically in mid to late adulthood, who neither have any family history of ataxia nor any acquired cause. The main idiopathic ataxia to exclude in such patients is the cerebellar form of multiple system atrophy (MSA), an α-synucleinopathy that can either present as parkinsonian-predominant (MSA-P) or cerebellar ataxia-predominant forms (MSA-C).^1^ Many patients with MSA, and especially MSA-C, have comparable clinical features to ILOCA that develop in mid-life, namely cerebellar ataxia, with cerebellar atrophy on brain imaging.^3^ The variable features also include tremor, dysarthria and dysphagia, and more rarely cognitive impairment with executive dysfunction. However, MSA is invariably associated with dysautonomia and is more rapidly progressive to death. MSA does not have any established genetic basis, although several genes have been suggested including repeat expansions in genes known to cause familial forms of ataxia.^2^ Given the strong association between expansions in SCA27B and ILOCA, we considered it important to test for an association between pathologic expansions in this gene and MSA in an ongoing, longitudinal study from South Korea.^3,4^

## RESULTS

The MSA Coalition Collaborative Core Network was developed to address its genetic etiology by assembling samples and genome sequences from well-characterized patients in ethnically diverse populations.^5^ Here we focus on a South Korean cohort^4^ that includes 199 individuals (91 male, 108 female) with a mean age at an examination of 58.2 ± 8.3 (median 58; range 34-79). All are ‘probable MSA’ according to the 2nd consensus criteria for diagnosing MSA.^6^ Our cohort includes 122 patients with MSA-C and 77 MSA patients with MSA-P. There are 80 MSA-C patients without parkinsonism, 42 MSA-C with mild parkinsonism, 62 MSA-P individuals without ataxia, and 15 MSA-P patients with mild ataxia. A total of 137 patients had cerebellar ataxia in their diagnosis (Table 1).

**Table 1.** Clinical characteristics of Korean patients with MSA and control series.

| Diagnosis | Count | Female | Male | Age, at diagnosis<br>(years) | Cerebellar ataxia | Parkinsonism |
| --- | --- | --- | --- | --- | --- | --- |
| MSA-C | 122 | 62 | 60 | 59.7 ± 9.4 | 122 | 42 |
| MSA-P | 77 | 46 | 31 | 57.4 ± 7.4 | 15 | 77 |
| Controls | 196 | 117 | 79 | 64.4 ± 9.0 | - | - |
| KGP controls<sup>9</sup> | 1,048 | - | - | - | - | - |

Whole genome sequencing has been completed for this entire MSA cohort. The minimum mean sequencing depth for all samples was 35x. All non-synonymous single nucleotide variants (SNVs) and small insertions and deletions (indels) in *FGF14* were investigated. None were consistent with SCA27A^7^, and no variants have a known pathogenic or likely pathogenic annotation in Clinvar.^8^ All genomic variability observed in *FGF14* in MSA was also examined in jointly called WGS data from the Korean Genome Project (KGP_controls_ n=1,048).^9^ One MSA-C patient has a rare coding variant observed in exon 5 c.707C>T (p.Ala326Val; NC_000013.11:g.101722883G>A; MAF KGP_controls_= 0.0019, gnomAD_EastAsia_=0.0002), whereas a total of ten patients had rare indel variants of uncertain significance (VUS) within the 3’UTR, namely three MSA-P patients and one MSA-C patient with c.*971_*972dup (NC_000013.11: g.101721858_101721859insTT; MAF KGP_controls_=0.021, gnomAD_EastAsia_=0.008), five MSA-C patients with c.*972del (NC_000013.11:g.101721859del; MAF KGP_controls_=0.024, gnomAD_EastAsia_=0.008) and one MSA-C patient with c.*1609_*1612del (NC_000013.11:g.101721219_101721222del; MAF KGP_controls_=0.0005, gnomAD_EastAsia_=0.0000) (Table 2).

**Table 2:** ***FGF14* non-synonymous SNVs and indels, among patients**

| ***FGF14* genomic variant**<br>**(NC_000013.11)** | **c.DNA**<br>**(protein change)** | **Clinvar** | **MSA-**<br>**C** | **MSA-**<br>**P** | **MSA**<br>**MAF** | **KGP**<br>**MAF** |
| --- | --- | --- | --- | --- | --- | --- |
| g.101722883G>A | c.707C>T (p.A236V) | NA | 1 |  | 0.0025 | 0.0019 |
| g.101721858_101721859insTT | c.*971_*972dup | VUS<sup>*</sup> | 1 | 3 | 0.010 | 0.021 |
| g.101721859del | c.*972del | VUS<sup>*</sup> | 5 |  | 0.013 | 0.024 |
| g.101721219_101721222del | c.*1609_*1612del | VUS<sup>*</sup> | 1 |  | 0.0025 | 0.0005 |

\*Variant of Uncertain Significance (VUS); NA= Not Available.

Subsequently, using the EH algorithm,^10^ we interrogated all MSA genomes for the *FGF14* 5’-flanking (GAA)_n_ repeat which is reported to be pathogenic when expanded.^11^ The (GAA)_n_ triplet repeat number was 17.8 ± 16.2 SD, range 9-111, n=398 alleles in Korean MSA patients, versus 17.9 ± 17.2 SD, range 0-162, n=2096 alleles in KGP controls. The most frequent (GAA)_n_ repeat number per allele was 10 (27.4%), followed by 9 (21.9%), 11 (8.8%), 24 (8.3%), 18 (4.8%), 17 (3.8%), 22 (3.3%), 25 (2.5%), 26 (2.5%) and others (16.7%). High-throughput sequencing in 1000 Genome samples highlights other motifs, most especially (GAAGAAGAAGAAGCA)- and (GAAGCA)-containing repeats in East Asian samples. Thus all known combinations were included in our EH search catalog (Supplementary table 1). However, comparable results and minor allele frequencies (MAF) were predicted for all motifs in cases and controls (Supplementary figure 1).

We sought to validate allele sizes based on EH results with PCR-based amplicon length analysis. The *FGF14* (GAA)_n_ genomic interval is 245,113bp 5’ of the start of the gene (NCBI reference NG_008317.3). Here, the wild type reference sequence has 50 pure GAA repeats spanning 150 bp. PCR primers were designed to span this interval to give a 315 bp product of which 165 bp is flanking sequence. However, PCR conditions were optimized to ensure positive controls with *FGF14* (GAA)_n_ expansions in the pathologic range would also be amplified.^11^ *FGF14* (GAA)_n_ locus PCR was subsequently performed in 199 MSA patients and 196 ethnically matched control participants (Fig. 1, Supp Fig. 2-3). Product sizes were estimated by gel migration and overall the distributions in control individuals (mean= 37±SD 57 bp, min-max:10-350, n=392) and MSA (mean= 44±SD 68 bp, min-max:10-500, n=344) were not significantly different (two sample t-test assuming equal variances, t=1.65, p>0.07). The majority of allele sizes in both groups were ≤50 bp including 89.2% (350/392) of controls and 86% (296/344) of MSA. However, some expanded alleles ≥150 bp were found in 6.9% (27/392) of controls (mean=224±SD 64 bp, range 150-350 bp) and 13.4% (46/344) of MSA (mean=195±SD 80 bp, range 150-500 bp).

**Figure 1.**
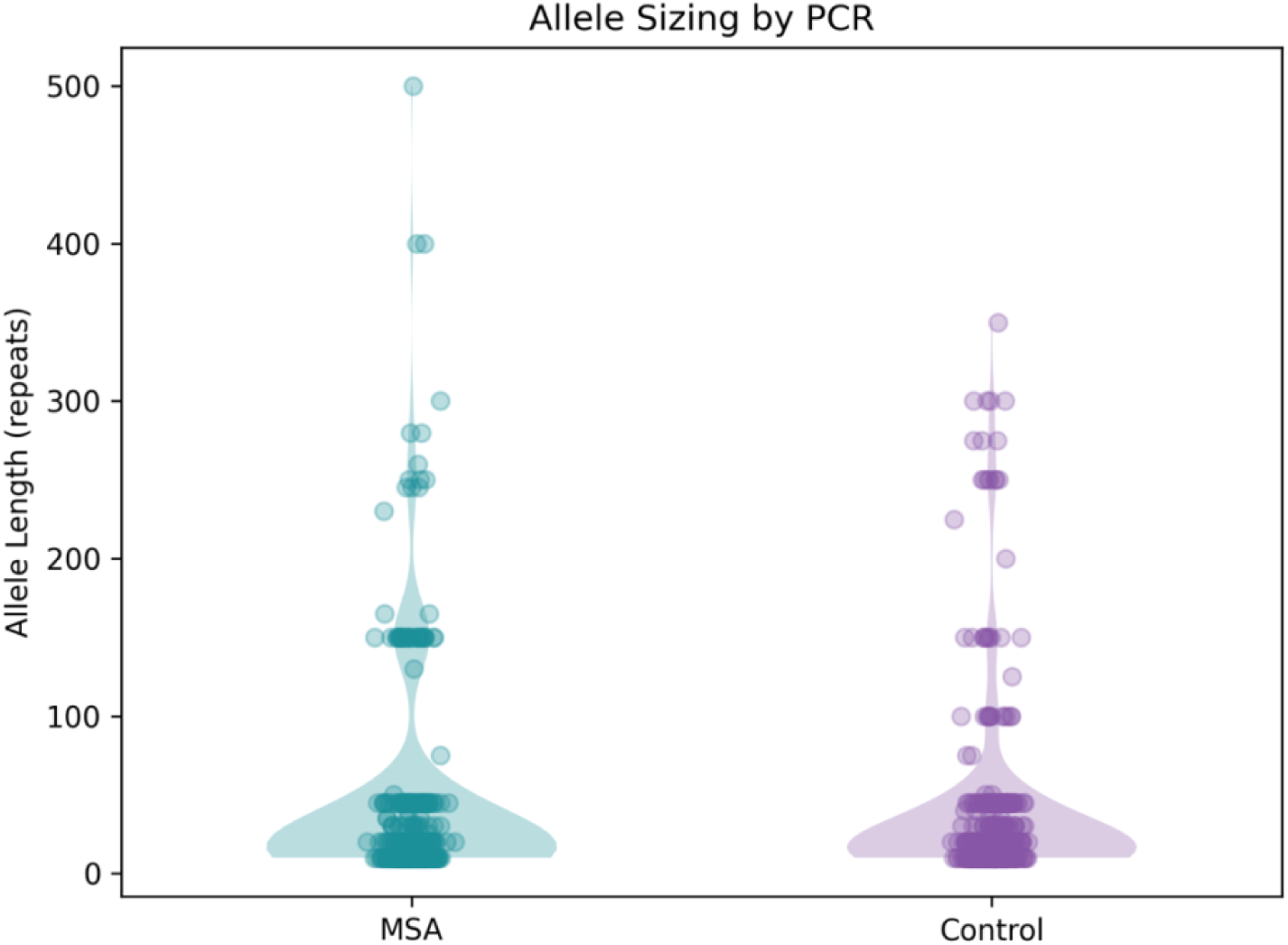
*FGF14* alleles size distributions from amplicon PCR and gel electrophoresis.

**Figure 2.**
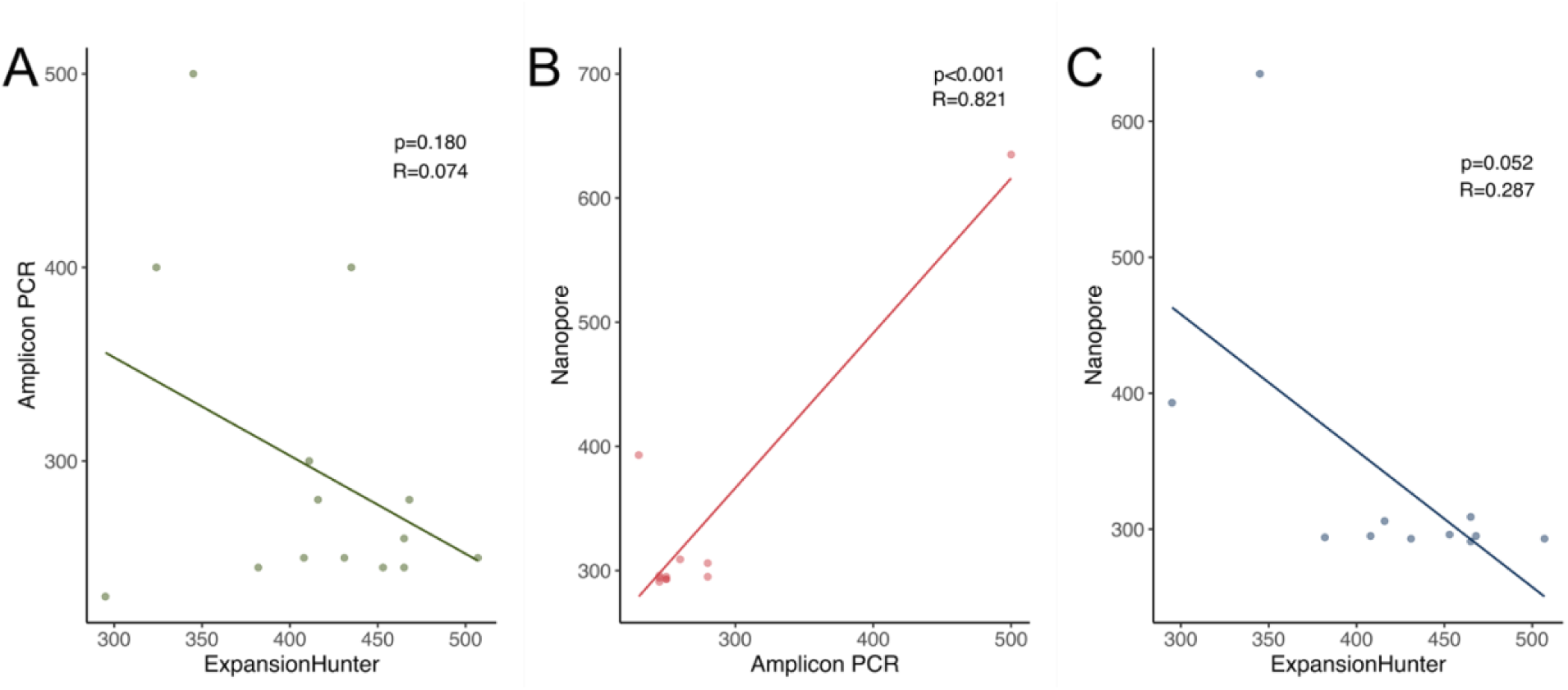
Size correlation between ExpansionHunter, amplicon PCR with gel sizing and Nanopore sequencing. A linear regression model was used for statistical analysis. Amplicon PCR with gel sizing vs EH: p=0.180, R^2^=0.07 (n=15); EH vs Nanopore: p=0.052, R^2^=0.29 (n=12); Amplicon PCR with gel sizing vs Nanopore: p=0.753×10^-4^, R^2^=0.82 (n=12). Legend: EH = ExpansionHunter; Nanopore = Nanopore sequencing.

As amplicon PCR with gel sizing and EH results were inconsistent we used linear regression to assess their relationship (p=0.18, R^2^=0.07)(Figure 2A, Supplementary figure 2 and 3). A subset of expanded alleles (n=11) was also interrogated using long-read sequencing on an Oxford Nanopore PromethION. The median quality of reads (phred score) was q=13.4, and the median read length (including both alleles) was 0.934 kb. The mean read coverage was 63,411X. The results of Nanopore sequencing were more consistent with amplicon PCR gel sizing results (p=0.75×10^−4^, R^2^=0.82) (Figure 2B), and it was possible to detect a complex repeat interruption pattern in 10/11 patients with the long expansion (GCAGAAGAAGAAGAA)_n_(GCAGAA)_n_ (GAA)_n_(GAG) (Table 3). Long-read sequencing supports PCR gel sizing results, showing similar (GAA)_n_ repeat expansion sizes and the length of those repeats before variable interruptions differed between the patients. However, long-read sequencing does not correlate with EH for the (GAA)_n_ expansion size (p=0.052, R^2^=0.29) (Figure 2C).

**Table 3.**
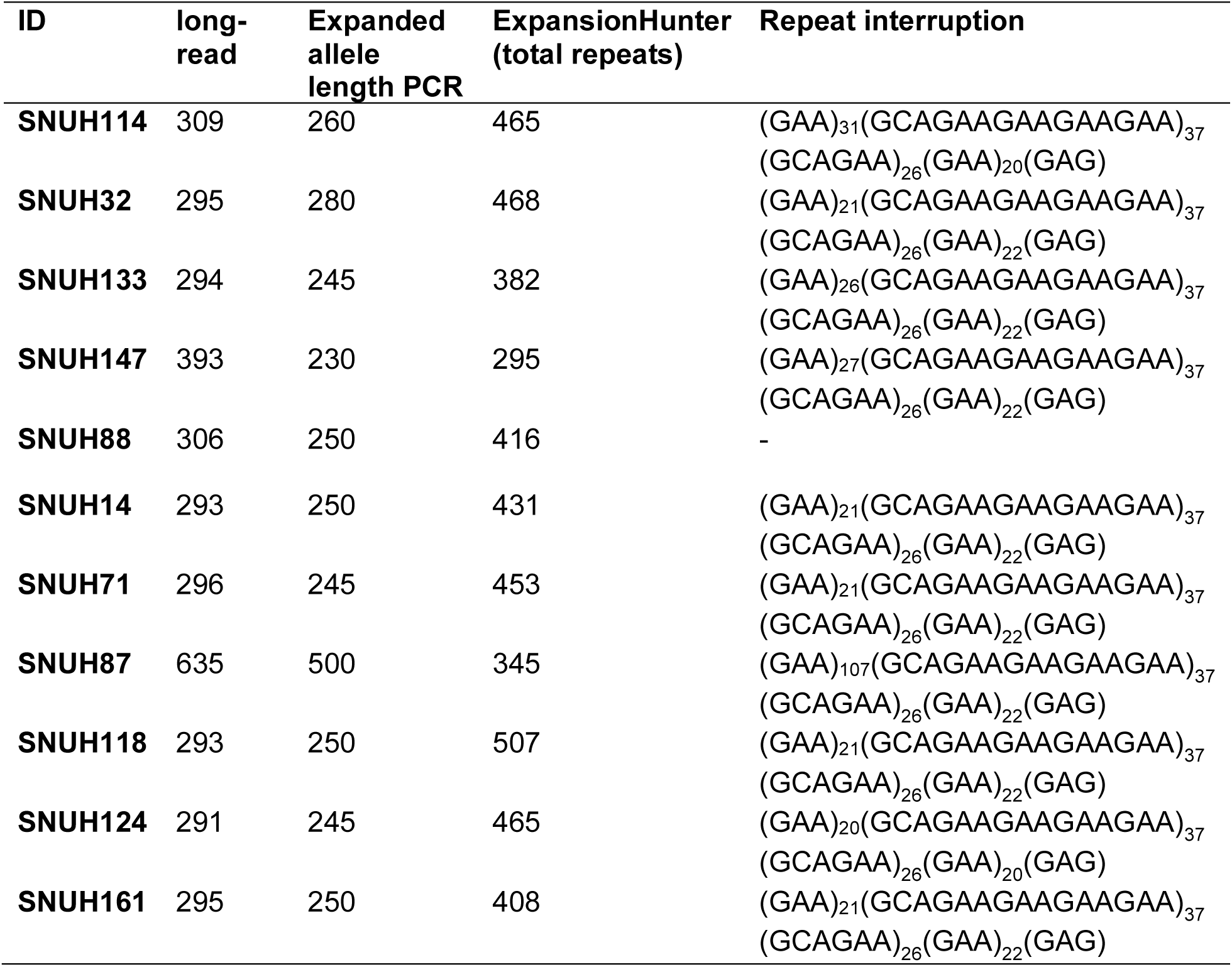
Repeat interruptions identified in expanded alleles via long-read sequencing.

## DISCUSSION

*FGF14* haploinsufficiency and loss-of-function clearly contributes to spinocerebellar ataxia type 27A (SCA27A) and SCA27B^9,13–15^ but there is less evidence that genetic variability at the *FGF14* locus contributes to MSA. Our results exclude *FGF14* (GAA)_n_ pathogenic repeat expansions, and rare *FGF14* coding variants, although we observe a higher frequency of intermediate expanded alleles in patients (13.4%) compared to controls (6.9%). While our sample size is too small for significance further meta-analysis may reveal an association with MSA susceptibility. Amplicon PCR gel sizing and long-read sequencing were highly correlated, but the former is not a precise tool for analyzing exact repeat lengths nor repeat interruptions (Table 3). Unfortunately, EH was not able to accurately annotate long, complex expanded repeats, including *FGF14* (GAA)_n_ (Fig 2C). This was demonstrated by the difference between EH and PCR or long-read results (Table 3). Thus, for future genetic testing, we recommend using amplicon PCR analysis to highlight potential (GAA)_n_ repeat expansions >250 bp, with subsequent confirmation by long read sequencing to most accurately determine the repeat size and composition, and an individual’s susceptibility to disease.

In this sample, in which the MSA-C subtype was more prevalent than MSA-P, genetic variability within the *FGF14* locus, including the (GAA)_n_ repeat, does not appear to contribute to disease. Nevertheless, an international meta-analysis on the role of *FGF14* in ataxia syndromes would be worthwhile, with appropriate standards and methods. Founder effects are often identified in repeat expansion diseases,^18^ with predominant haplotypes associated with expanded alleles from specific geographic disease clusters.^16^ A disease-associated *FGF14* haplotype has been observed in French-Canadian patients, and *FGF14* (GAA)_n_ expansions appear to be a more frequent stochastic event in sporadic ataxias.^15^ As *FGF14* (GAA)_n_ late-onset ataxia may also be responsive to 4-aminopyridine (4-AP), a genetic diagnosis may inform treatment options.^17^

To better understand the genetic architecture underlying MSA, we continue to sequence samples and compile data from patients, and we jointly report on their genome variability, including similar analysis of *de novo* repeats. We host the data in the computational genome analysis platform, CGAP (https://cgap.hms.harvard.edu) which was chosen for the storage, visualization, and analysis of data from Mission MSA. CGAP implements a variety of bioinformatics pipelines and performs a wide range of analyses on genome data, and would welcome additional members and participation.

## METHODS

### Study Overview

Patients were enrolled in the Seoul National University Hospital (SNUH) outpatient clinic setting from December 2012 to April 2021. The eligibility criteria for the study included patients with ‘probable MSA’ according to the 2nd consensus criteria. Following enrollment, 10ml peripheral blood was collected for biobanking, including DNA extraction using standard methods. A detailed medical and family history was collected during the patient interview and recorded in the electronic medical record (EMR). Family history was obtained for any potentially related neurologic disease. To obtain a full medical history of participants, their EMR was also reviewed.

### Institutional review board

This study was approved by an institutional review board (IRB) protocol of the SNUH: H-1601-048-733 and all participants provided written informed consent. To protect privacy, each participant is assigned an anonymous identification number. In addition, this study protocol was reviewed and IRB approved by the University of Florida #IRB202000632, which enables whole genome sequencing in brain health and disease. Additional anonymized genomic data was made available from healthy Korean volunteers participating in the Korean Genome Project (KGPcontrols).^9^

### Whole genome sequencing (WGS)

WGS was performed by the Clinical & Translational Genomics Program at the University of Florida. Samples and data are processed and stored according to HIPAA compliance requirements following CAP guidelines^12^ and CLIA standards for quality and competence^13^ at UF Health Medical Laboratories. Testing has been benchmarked using the National Institute of Standards and Technology ‘Genome in a Bottle’ Consortium standards (HG002 son, HG003 & HG004 parental genomes).^14^ WGS precision metrics have been validated from 5-30× depth for the entire genome and results are concordant with the NIST/PrecisionFDA data “truth sets”^15–18^ with > 95.2% analytical sensitivity and > 97.3% precision.

Genomic DNA was extracted from blood using QIAamp™ protocols and quantified by fluorescence on an Invitrogen Qubit Fluorometer. Individually-indexed genomic libraries were prepared using dual unique indexes from ∼200ng DNA/individual (New England Biolabs NEBNext^®^ Ultra^TM^ II DNA Library Prep Kit for Illumina^®^). Genome library quality and quantity were confirmed by automated electrophoresis on an Agilent 2100 bioanalyzer and by qPCR. Individual libraries were then normalized and pooled in equimolar ratios for 2× 150 bp paired-end sequencing at 35× depth on an Illumina NovaSeq 6000. To expedite and enable innovation, speed, data and code sharing, and ensure security and PHI/HIPAA compliance, we maintain a containerized compute environment in Amazon Elastic Compute Cloud (AWS EC2) and a versioned bioinformatic pipeline for genome variant calling. In brief, fastQ reads are aligned to the human genome (GRCh38/hg38) with ‘versioned’ open-source software including TrimmomaticPE,^19^ FastQC,^20^ and MultiQC^21^. Computation is orchestrated by Nextflow (more detail is available on request). Initial quality control (QC) reports are generated and examined for all individual samples and batched runs, including general statistics on WGS coverage per sample, mapping quality, and the proportion of reads mapped. These data quantify sequence read counts (unique, duplicate, and overrepresented %) and quality (Phred scores across reads, per sequence quality scores, length distributions, GC content, and ‘N’ scores). Data passing QC is transferred to CGAP via AWS protocols for secondary QC and bioinformatic analyses.

### Amplicon PCR

Published methods were used to amplify the FGF14 (GAA) locus,^11^ the size of the alleles generated was visually sized on 2% agarose by gel electrophoresis. *FGF14* can be found in NCBI reference sequence NG_008317.3 and contains the genomic interval Chr13: 102,161,440 – 102,161,888 (GRCh38) that is pathologically expanded at 245,113bp 5’ of the start of the gene. This NCBI reference sequence is wild type for a 150 bp region of 50 pure GAA repeats. PCR primers 5’-AGCAATCGTCAGTCAGTGTAAGC (FGF14 Forward) and 5’-CAGTTCCTGCCCACATAGAGC (FGF14 Reverse) span this interval to give a 315bp product.

### Long-read sequencing (LRS)

Expanded samples were selected to characterize the repeat domain of *FGF14*. Amplicon PCR products were visualized using the 4200 Tapestation and underwent purification using SPRI beads to isolate a targeted region of >500bp. The Native 24 Barcoding Kit (SQK-NBD114.24) was used for multiplexing the PCR products, and 1 µg of amplified PCR product was used as input. The following sequencing adapter ligation was performed with the SQK-NBD114.24 (ONT). The final product was then loaded on R10 PromethION flow cells on a P24. The input for the library preparation was 200 fmol of DNA per sample. Base-calling was performed with the high accuracy model of Guppy version 7.1.4. Only reads with a base-calling accuracy of over 90% were included. To analyze the quality of the reads, the software Nanostat (version 1.5.0) was used. For the alignment to the reference sequence, Minimap2 (version 2.22) was used.^22^ The handling of SAM/BAM files as well as the calculation of the coverage was performed with Samtools (version 1.15).^23^ Only reads with an alignment length over 1kb were included in the analysis. Finally, the detection of the hexanucleotide repeats was performed using the “Noise-Cancelling Repeat Finder” (NCRF, version 1.01.02).^24^ For the calculation of repeat length, the median of all reads was used, as previously described.^25^ The exploration of repeat interruptions was done with the NCRF alignment, as previously described.^26^

### Bioinformatic analysis

The bioinformatics analysis was performed using CGAP pipelines, and automatically executed in AWS cloud infrastructure using Tibanna^27^, an open-source software for pipeline execution management. The pipeline follows Genome Analysis Toolkit (GATK) best practices^28^, and for germline SNVs and indels, runs GATK variant calling algorithms with increased sensitivity and two additional inheritance-mode calling algorithms (novoCaller^29^ and comHet^30^) to refine and prioritize rare events in GATK standard calls.

Paired-end FASTQ files were aligned to GRCh38 reference genome using bwa-mem, followed by cleanup to remove duplicate reads (MarkDuplicates) and recalibration of base quality scores (BaseRecalibrator and ApplyBQSR). Variants were called for each BAM file using HaplotypeCaller and jointly genotyped (CombineGVCFs and GenotypeGVCFs) within the family where family members were available. The raw calls were then processed to split multi-allelic variants, re-align indels, and remove variants that did not reach a sufficient read depth coverage. Variants were annotated using the Ensembl Variant Effect Predictor (VEP) together with multiple external data sources (i.e., dbNSFP, MaxEnt, ClinVar, SpliceAI, gnomAD V2 & V3, CADD, phastCons, phyloP, GERP, SIFT, PolyPhen2, REVEL, PrimateAI) and underwent a series of filtering steps to remove intergenic variants, non-functional variants, and common variants in the population with a Minor Allele Frequency (MAF) > 1%. Finally, the remaining calls were refined by running the inheritance-mode calling algorithms novoCaller and comHet to detect *de-novo* and compound heterozygous variants for probands with sequenced family members.

*FGF14* non-synonymous variants were selected from the annotated VCF file generated by the pipeline. Selection criteria included variants that had a moderate or high impact with MAF < 1% in the East Asian population, or a ClinVar annotation of “pathogenic”, “likely pathogenic”, or “VUS”.

Known pathogenic repeat expansions were screened with the ExpansionHunter algorithm^10^ (v.5.0.0) using default parameters. A catalog of pre-defined pathogenic repeats consisting of repetitions of short sequence units (1-6 bp) was used for screening, with the deep intronic *FGF14* (GAA)_n_ repeat expansion introduced at chr13:102161575-102161724. The number of *FGF14* (GAA) repeats on each allele was estimated using information from reads that span, flank, and are fully contained in each repeat.

Figure 1B and Figure 2 are generated with standard Python packages, including Matplotlib and Scikit-learn for linear regression calculations.

## Data Availability

All data produced in the present study are available upon reasonable request to the authors.

## Acknowledgments

We thank all patients and families who participated in this study. We thank Dr. Paul Lockhart for a kind gift of amplicons with *FGF14* (GAA)_n_ expansions in the pathologic range. We thank previous members of the Laboratory of Neurogenetics and Neuroscience for their technical support, and the Interdisciplinary Center for Biotechnology Research (ICBR) at the University of Florida. This work was made possible by an Award from Mission Multiple System Atrophy. CGAP is also supported by the Blavatnik Clinical Pilot Award to Drs. Peter Park and Shamil Sunyaev, The Chan Zuckerberg Initiative EOSS5 grant 2022-309591 (an advised fund of the Silicon Valley Community Foundation) to D.V., The Department of Biomedical Informatics and The Park Lab at Harvard Medical School. MJF was supported through the Clinical & Translational Genomics Institute.

---

**Supplementary table 1.**
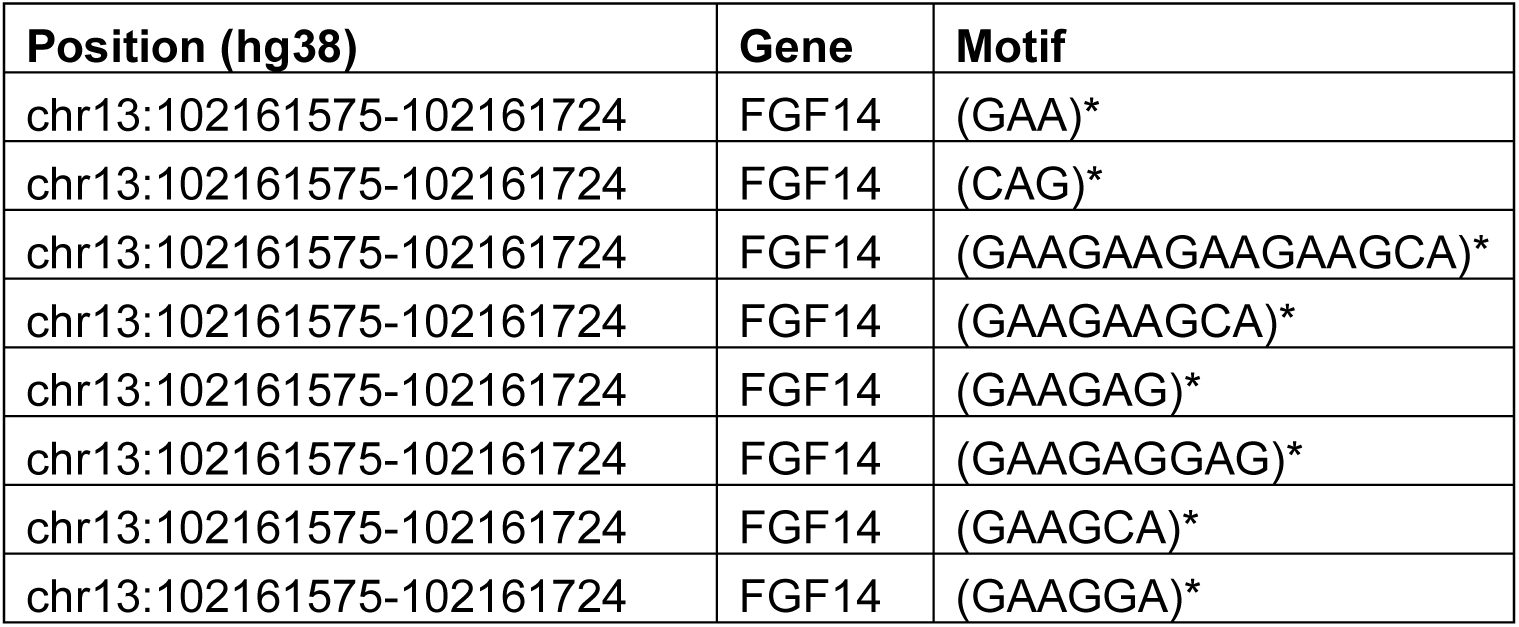
*FGF14* repeat motif catalogue examined with Expansion Hunter.

**Supplementary Figure 1.**
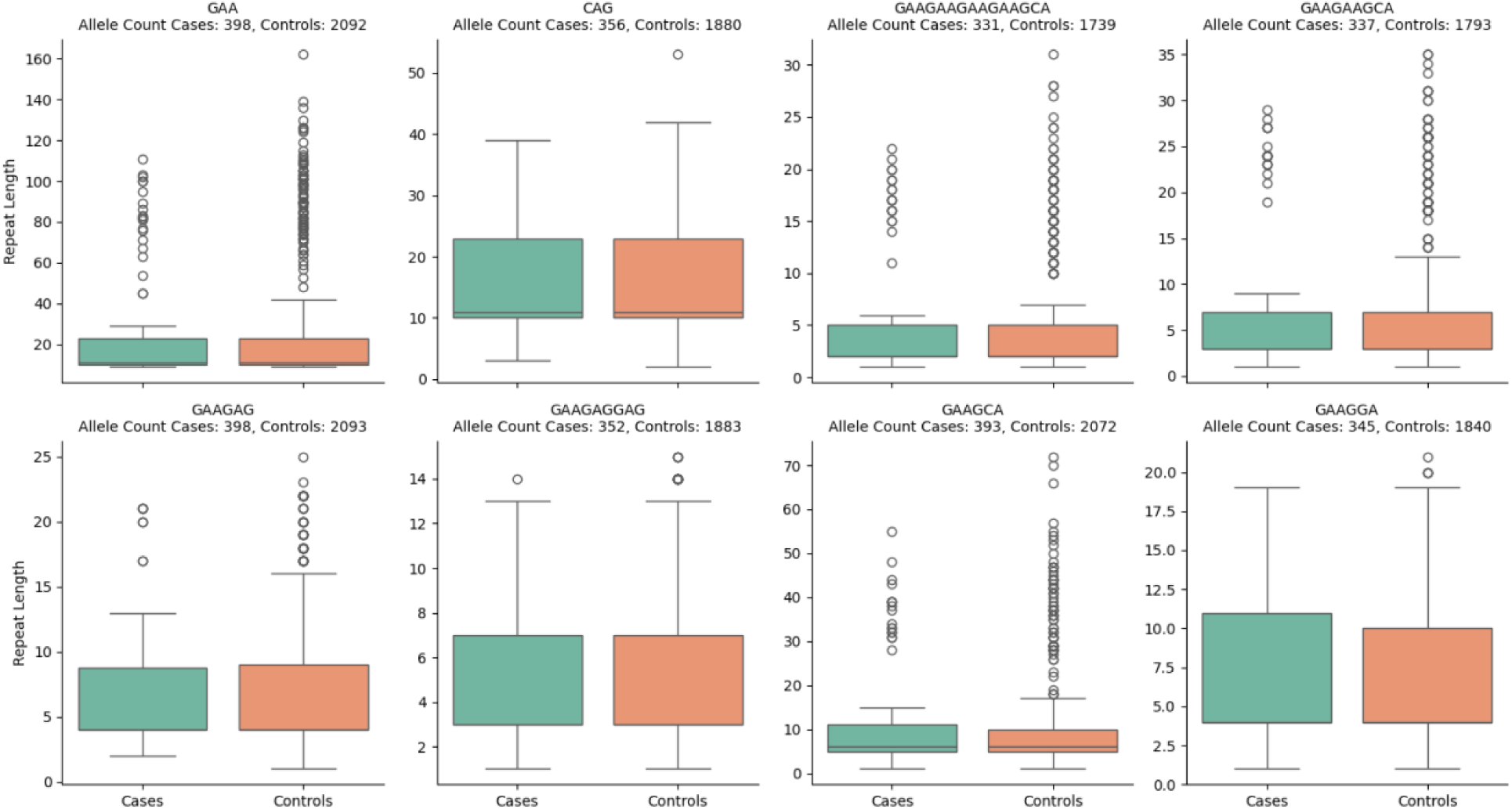
Analysis of separate *FGF14* repeat motifs and sizes, in MSA cases and controls, estimated with Expansion Hunter.

**Supplementary Figure 2.**
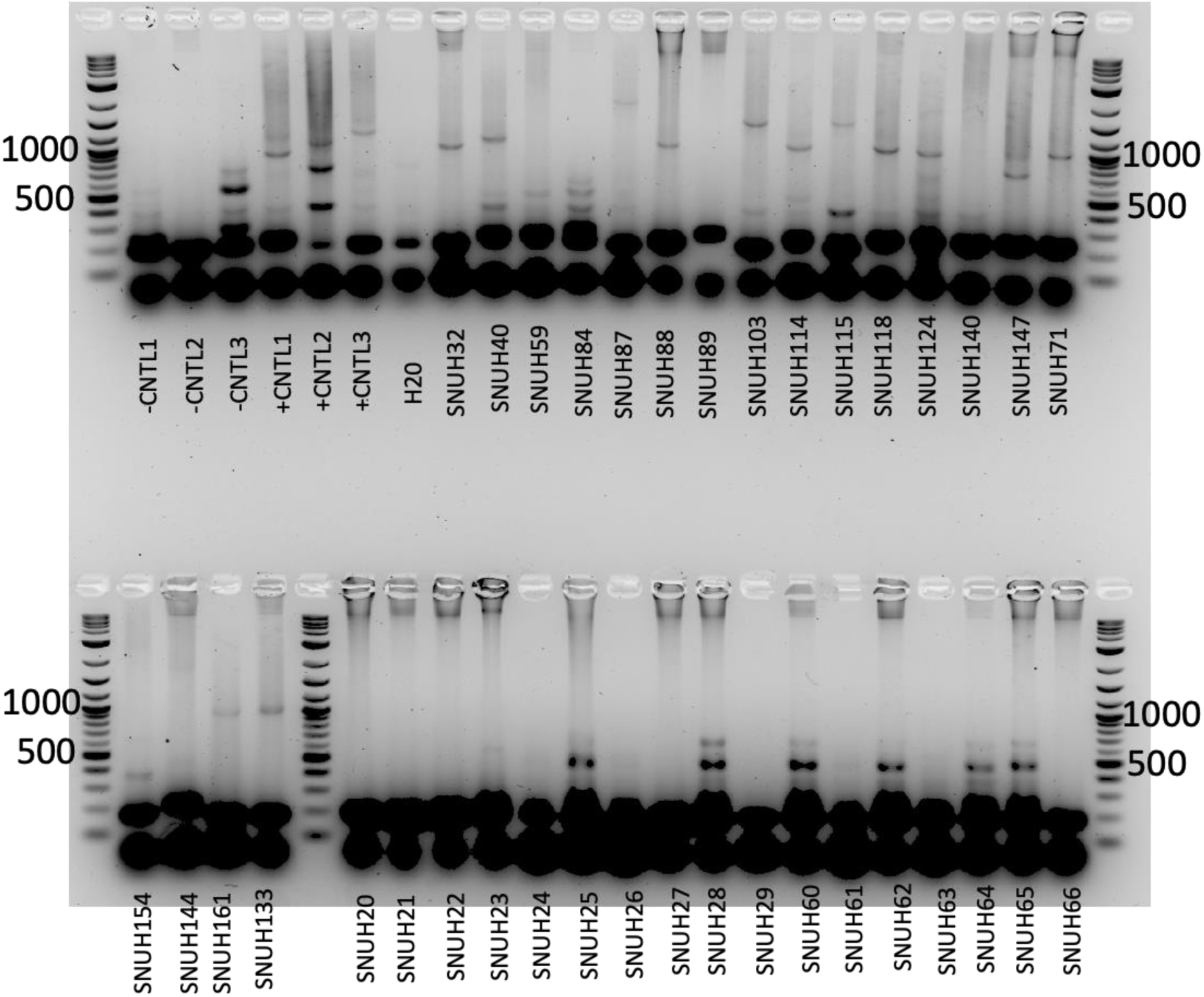
*FGF14* amplicons with gel sizing. *FGF14* negative and positive controls have the following (GAA)n repeats, respectively from top left: -CNTL1 (13,22); -CNTL2 (13,22); -CNTL3 (13,45); +CNTL1 (20,245), +CNTL2 (185,289), CNTL3 (13,349).

**Supplementary Figure 3.**
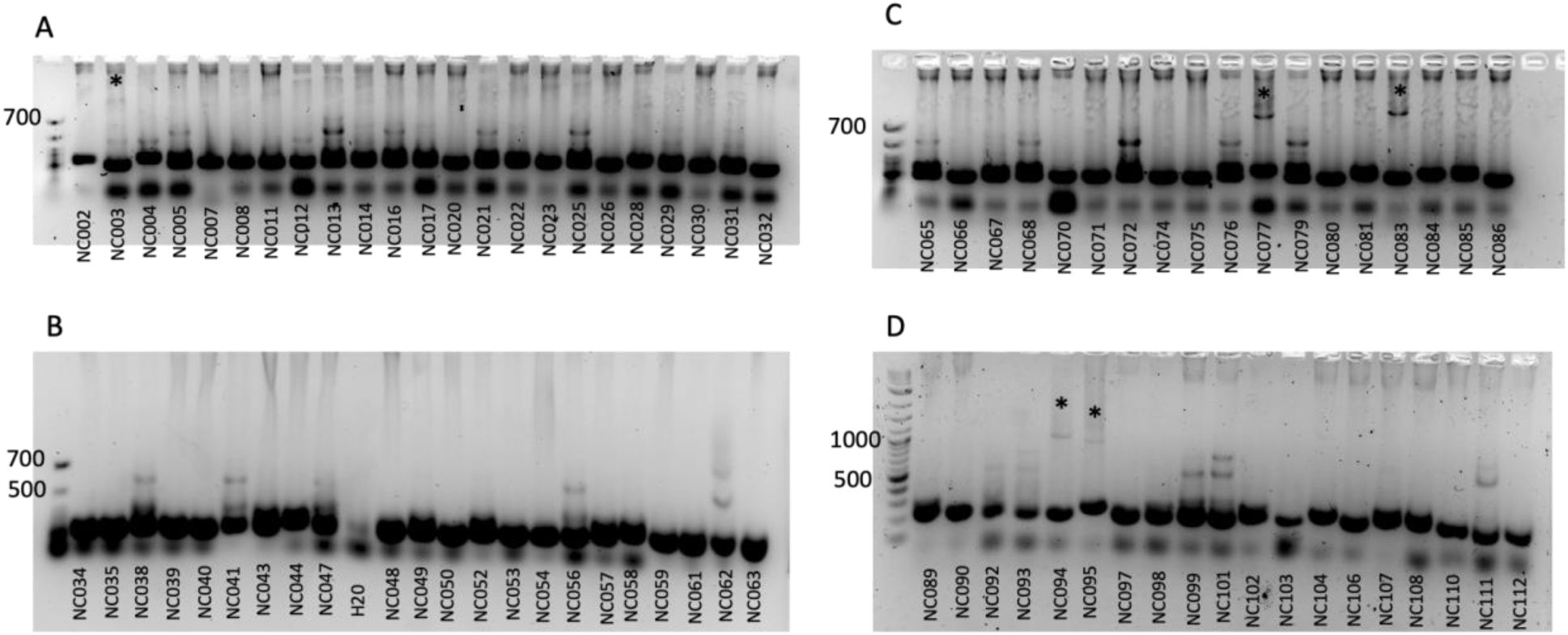
Korean controls. (A-D) Full gels of PCR amplicons for Korean controls (NC). * symbolizes an FGF14 amplicon that is greater than 200. (A) NC002 to NC032, (B) NC034 to NC063, (C) NC065 to NC086, (D) NC089 to NC112.

## Notes

### Competing Interest Statement

The authors have declared no competing interest.

### Author Declarations

Institutional review board of the Seoul National University Hospital gave ethical approval for this work. Institutional review board of the University of Florida gave ethical approval for this work.

